# Do Classical Speed-Accuracy Trade-Off Models Generalize to Lower Limb Movements?

**DOI:** 10.64898/2026.10.07.26364578

**Authors:** Paola Di Florio, Asli I. Karamanlargil, Wiebren Zijlstra, Deepak K. Ravi, Olivier Lambercy, Roger Gassert, Eleftheria Giannouli

**Affiliations:** Rehabilitation Engineering Laboratory, Department of Health Sciences and Technology, ETH Zurich, Switzerland; Department of Biomedical and Neuromotor Sciences, University of Bologna, Italy; Institute of Movement and Sport Gerontology, German Sport University Cologne, Cologne, Germany; Laboratory for Movement Biomechanics, Department of Health Sciences and Technology, ETH Zurich, Germany

**Keywords:** Speed-accuracy trade-off, Fitts’ law, Stepping task, Anticipatory postural adjustments, Reaction time, Foot placement accuracy, Postural control

## Abstract

Targeted stepping requires coordinating limb transport, preparatory postural adjustments, and accurate foot placement while maintaining balance under competing speed and accuracy demands. Although Fitts’ law robustly describes speed-accuracy trade-offs (SATs) in upper-limb movements, its applicability to whole-body stepping remains unclear due to additional postural and visuomotor constraints. This study investigates how instructional emphasis and spatial demands modulate SAT regulation and pre-step postural dynamics during stepping. Twenty-four healthy young adults will perform a multidirectional stepping task across two directions (0° and 45°) and two distances (45% and 80% of leg length) under three instruction conditions: Fast, Accurate, and Fast & Accurate. Kinematic and force-plate data will quantify movement time, foot-placement error, and pre-step center-of-pressure dynamics. We hypothesize that instructional emphasis and increased postural demands will systematically alter stepping strategies and produce deviations from classical Fitts-type scaling, particularly during diagonal stepping.

## Introduction

Effective execution of a targeted step requires identifying a stepping target, organizing a center of pressure (COP) shift to redistribute body weight, producing sufficient force to initiate movement, and placing the foot with adequate accuracy (Day and Bancroft, 2018). Disruption of any of these processes can compromise step timing or stability, thereby increasing fall risk (Okubo et al., 2021). Because stepping must be both rapid and precise to maintain balance, its control inherently involves a trade-off between speed and accuracy. Targeted stepping therefore provides a well-suited context in which to examine how the nervous system coordinates these competing demands.

A fundamental principle of motor behavior governing the regulation of these competing demands is the speed-accuracy trade-off (SAT) (Schmidt et al., 2019). When individuals attempt to move faster, errors typically increase; conversely, prioritizing accuracy prolongs movement time (Bogacz et al., 2010; Heitz, 2014). This relationship has been extensively studied in upper-limb targeted movements and formalized in Fitts’ law (Fitts, 1954).

Fitts law describes a linear relationship between movement time (MT) and the index of difficulty (*ID*) (eq. 1)

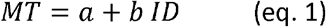

Where *a* is the intercept, reflecting baseline movement time, i.e., the minimum time it takes for a person to initiate and complete a movement when task difficult approaches zero, and *b* is the slope, reflecting sensitivity to task difficulty. In its original formula, the ID is expressed as:

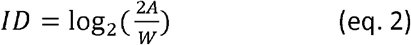

where *A* is the movement amplitude (target distance) and *W* is the target width.

Although extensively validated for upper-limb actions (Plamondon & Alimi, 1997; Schmidt et al., 2019), the extension of Fitts’ law to lower-limb behavior is not straightforward due to additional postural control requirements. Unlike the upper limb, the lower extremity is functionally coupled to whole-body balance, requiring preparatory and reactive control of the center of mass during movement initiation. Consequently, step execution depends on a preparatory COP shift prior to foot-off, introducing biomechanical constraints absent in upper-limb tasks.

Evidence from locomotor targeted tasks suggests that speed-accuracy trade-offs extend to whole-body actions, such as running toward spatial targets, supporting the idea that Fitts-like principles may generalize beyond discrete upper-limb movements (Bradshaw & Sparrow, 2000). Consistently, lower-limb motor control studies report that movement time generally increases with task difficulty in a broadly Fitts-like manner, although systematic deviations from classical Fitts’ law are frequently observed. These deviations indicate that a single index of difficulty may be insufficient to fully capture performance when postural and whole-body constraints are involved. In particular, foot-pointing and other constrained lower-limb targeted movement studies show that foot-placement accuracy-related measures scale with task difficulty, whereas movement time does not consistently follow classical Fitts’ law scaling, likely reflecting that accuracy can be preserved given sufficient time, while movement speed is bounded by biomechanical constraints (Luchies et al., 2002; Duarte & Latash, 2007). Similarly, in tasks combining lower-limb targeted movements with postural regulation, COP-related variables tend to scale with task demands, whereas temporal measures often show weaker or inconsistent adherence to Fitts-type scaling, likely due to additional constraints imposed by whole-body stability requirements (Bertucco & Cesari, 2010; Bancroft & Day, 2016).

Taken together, these findings highlight a key distinction between tasks in which the lower limb operates primarily as an effector for endpoint control and those in which it is embedded within a full-body balance task. In contrast to foot-pointing paradigms, whole-body stepping tasks require the coordination of limb transport with dynamic postural control. This introduces an additional layer of constraint, as movement execution is tightly coupled to preparatory COP adjustments that are essential for maintaining stability during step initiation (Uemura et al., 2013).

A key characteristic of stepping is its directional dependence. In particular, diagonal stepping requires simultaneous control of forward progression and mediolateral weight transfer. This increases postural demands relative to purely sagittal stepping, as balance must be maintained while generating oblique center of mass trajectories. Empirical evidence from stepping studies shows that COP strategies are direction-specific, with systematic modulations in amplitude and timing of preparatory adjustments during lateral and diagonal stepping conditions (Inaba et al., 2020; Hayashi et al., 2016; Russo et al., 2021). Accordingly, multidirectional stepping represents a more demanding and ecologically valid context for investigating speed-accuracy regulation.

Beyond biomechanical constraints, task instructions strongly influence stepping behavior. Studies targeted stepping tasks demonstrate that prioritizing speed versus accuracy systematically alters movement execution and preparatory/anticipatory postural adjustments. For example, Uemura et al. (2013) showed that instructions significantly modulate step timing and COP control, revealing a clear instruction-dependent speed-accuracy trade-off. Similarly, Muijeres et al. (2023) demonstrated flexible adaptation of stepping performance across competing task-level goals, including speed, accuracy, and stability during stepping from quiet standing.

Importantly, most previous studies have used floor-projected targets that provide a direct spatial mapping between perception and action, such that the visual stimulus specifies the movement goal without requiring additional selection or transformation. In contrast, tasks in which movement targets are specified indirectly via visual cues require a visuomotor transformation, as individuals must interpret the cue and select the appropriate spatial goal prior to movement execution. This additional processing step introduces a decision component that may alter how spatial accuracy is encoded and controlled during movement planning, yet its influence on speed-accuracy trade-offs in stepping remains largely unexplored. Notably, such indirect visuomotor mappings are common in exergaming and virtual-reality-based (cognitive-motor) stepping exercises (Altorfer et al., 2021; Jäggi et al., 2023; Seinsche et al., 2023; Büttiker et al., 2024), where movements are guided by screen-based cues rather than directly specified spatial targets, further highlighting the ecological relevance of visuomotor transformations in stepping control.

If stepping performance reflects flexible regulation of speed and accuracy under combined biomechanical, postural, sensory, and cognitive constraints, instruction-based manipulations provide a direct way to examine these control strategies.

The present study therefore investigates whether instruction-driven prioritization of speed or accuracy produces stepping behavior consistent with Fitts’ law, or whether systematic deviations emerge due to postural demands and visuomotor transformations. This is tested using a multidirectional stepping paradigm including diagonal (45°) targets, implemented in a screen-based visual cueing paradigm and requiring both visuomotor transformation and dynamic balance control.

We hypothesize that:

1. Movement time will scale linearly with the Index of Difficulty across all instructional conditions; however, regression slopes and intercepts will differ as a function of instructional focus.
2. Speed-emphasis instructions will reduce movement time but increase foot-placement error, whereas accuracy-emphasis instructions will produce longer movement times and reduced spatial error.
3. Pre-step COP shift dynamics will be modulated by instruction, with speed-emphasis leading to shorter (in terms of duration) or smaller (in terms of amplitude) COP displacement and accuracy-emphasis leading to longer and possibly larger (in amplitude) preparatory shifts.
4. Deviations from classical Fitts-type scaling will increase in oblique (45°) stepping conditions, reflecting greater mediolateral balance demands compared to sagittal steps.
5. Instructional effects on speed-accuracy trade-offs will be amplified under screen-based visual cues presentation, due to the additional visuomotor transformation required compared to physically grounded targets.
6. Deviations from classical Fitts-type scaling will emerge when postural control variables are considered, indicating that movement time cannot be fully explained by a single index of difficulty.

## Methods

### Ethics Information

The study protocol was approved by the ethics committees of ETH Zurich (Nr: 26 ETHICS-053).

All interested participants will sign a written informed consent before any study procedures are carried out. No financial compensation will be provided for participation.

### Design

#### Inclusion & Exclusion Criteria

Twenty-four young and healthy adults (50% female) will be recruited via (i) announcements within the ETH Zurich community and the University of Zurich Alumni marketplace portal, (ii) posts in relevant Facebook groups, and (iii) posts by involved researchers via personal and institutional LinkedIn profiles. Inclusion criteria will be: aged between 18 and 40, ability to stand independently and perform multidirectional steps without assistive devices, speaking English. Exclusion criteria will be: neurological, musculoskeletal, or balance-related conditions that could affect stepping performance, and height exceeding 190 cm (due to technical constraints of the experimental plate).

#### Sociodemographic data

We will collect participants’ sex, age, level of education, profession, body height, leg length, and leg dominance through the question “If you would shoot a ball on a target, which leg would you use to shoot the ball?” (van Melick et al., 2017).

### Sampling Plan

#### Baseline Covariate Assessments

Prior to the experimental recordings, participants will complete two baseline assessments that can be included as covariates in the analyses to account for possible inter-individual differences in processing speed and balance performance. First, a simple reaction time test will be conducted using PsyToolkit (Stoet, 2010; Stoet, 2017), in which participants are instructed to press the spacebar of a keyboard as quickly as possible each time a cross appears on the screen. The *average response latency* will be computed as the mean reaction time across trials.

Second, a single-leg stance test (SLST) will be performed on the non-dominant foot, during which participants raise the dominant foot and attempt to maintain balance for 60 seconds (Nagymate et al., 2018) with the hands on the hips. During the SLST, participants will stand on a force plate to record the COP displacement, and retroreflective markers (Vicon Motion Systems Ltd, UK) will be placed on the feet to detect onset and offset of the test, as shown in figure 1.

**Figure 1.**
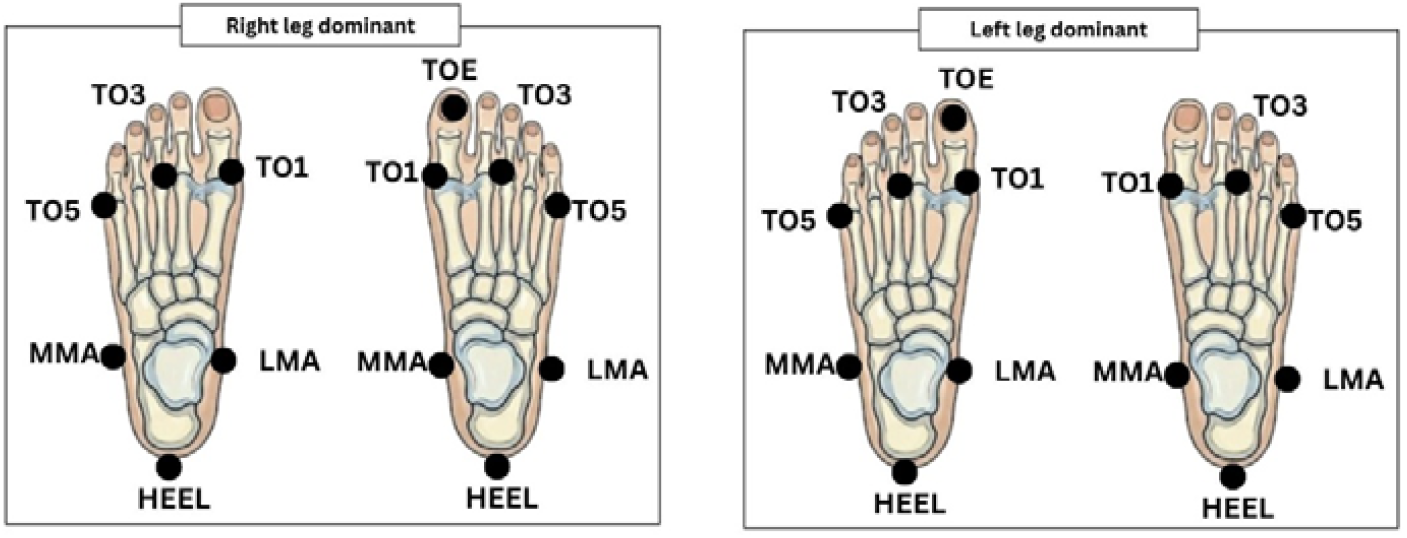
Markers protocol for each foot (HEEL: heel; LMA: lateral malleolus; MMA: medial malleolus; TO1: first metatarsal head; TO3: third metatarsal head; TO5: fifth metatarsal head; TOE: tip of the dominant toe).

#### Target Configuration and Calibration

Participants will stand on a force plate (9286B, Kristler, Switzerland) in a comfortable stance. Foot positions will be marked to ensure consistency across trials. Target locations will be defined using markers placed on the force plate, indicated by a cross with the size 2 × 2⍰cm square center and 4⍰cm arms. Near targets will be positioned at 45% of the participant’s leg length, and far targets at 80% of leg length, measured from the dominant toe during standing. Targets will be presented in two directions: 0° and 45° relative to the forward-facing stance. The calibration set-up is shown in Figure 2.

**Figure 2.**
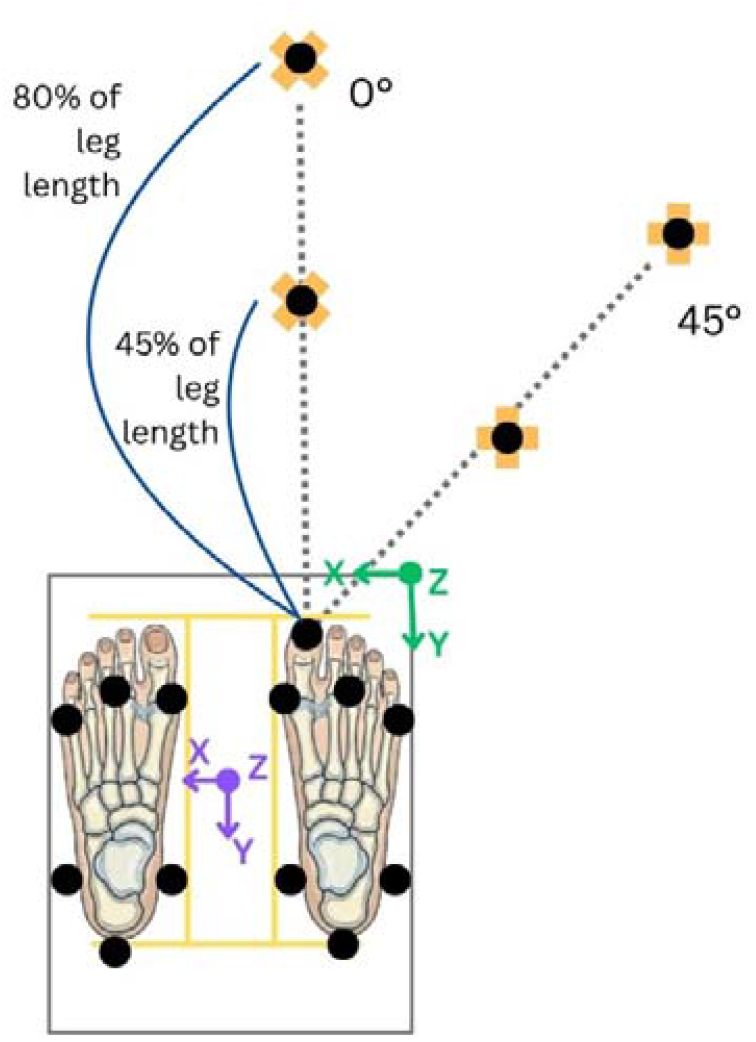
Initial calibration for mapping the positions of the markers, in the case of a right dominant foot. Black dots indicate retroreflective markers, while yellow crosses show the target positions. The origin of the stereophotogrammetric system is shown in green, while the origin of the force plate is shown in purple.

#### Experimental Task and Procedures

Participants will start each trial in a static neutral position, standing on the force plate with both feet placed in a self-selected comfortable stance. This stance will be determined during a calibration phase and marked on the force plate using adhesive tape to ensure consistent foot placement across all trials.

Visual stimuli will be presented on a screen positioned at floor level (resolution: 1920 × 1200 pixels). A target (red circle, 40-pixel diameter) will appear in one of four spatial locations (Fig. 2).

Prior to data collection, participants will complete three practice trials per target position to familiarize themselves with the task and minimize initial learning effects.

To assess speed-accuracy trade-offs, task demands will be manipulated via instructional emphasis. Participants will complete three instruction conditions:

- STANDARD: “As soon as a red circle appears on the screen, step as fast and as accurate as possible to the indicated position placing the toe on the center of the target cross and fully transfer your body weight onto the stepping foot raising the heel of the stance leg. When the circle turns green, return to the starting position at comfortable pace.”
- FAST: “As soon as a red circle appears on the screen, step as fast as possible to the indicated position placing the toe on the center of the target cross and fully transfer your body weight onto the stepping foot raising the heel of the stance leg. When the circle turns green, return to the starting position at comfortable pace.”
- ACCURATE: “As soon as a red circle appears on the screen, step as accurate as possible to the indicated position placing the toe on the center of the target cross and fully transfer your body weight onto the stepping foot raising the heel of the stance leg. When the circle turns green, return to the starting position at comfortable pace.”

Each block will last approximately 6 minutes. Targets (red and green circles) will be presented for 4 seconds, followed by a 1-second inter-trial interval. Participants will start with the STANDARD block, after which the order of FAST and ACCURATE blocks will be randomized and counterbalanced across participants.

Each block will include 40 trials (10 trials per target condition). If a trial is performed incorrectly (defined as adjusting the foot position after initial contact with the target), it will be repeated to ensure that each participant completes 10 valid trials per target, resulting in a total of 120 trials per participant.

During each trial, participants will step with the dominant foot towards the target, placing the toe on the target cross and transferring body weight onto the stepping foot while raising the heel of the stance leg. After reaching the target, participants will maintain the final position for 3 seconds without further adjustment before returning to the initial stance.

To minimize fatigue, participants will be given a 5-minute seated rest period between blocks. Target distances and directions were selected to systematically vary task difficulty. The near target (45% of leg length) was designed to require minimal trunk involvement and low mediolateral balance demands, whereas the far target (80% of leg length) approached the upper range of voluntary stepping capacity, increasing task difficulty without eliciting protective or multi-step responses. Leg length is defined as the distance between the greater trochanter and the lateral malleolus. Previous work has shown that larger step lengths increase swing duration, landing time, and balance recovery demands, whereas shorter steps impose minimal postural challenge (King et al., 2005).

Directional conditions further modulated task demands: the 0° direction represents a forward stepping movement, while the 45° direction toward the dominant side introduces lateral displacement, requiring greater hip abduction, trunk rotation, and postural control.

A summary of all task conditions, including instruction (STANDARD, FAST, ACCURATE), distance (near, far), and direction (0 °, 45 °), is provided in Table 1.

**Table 1.** Summary of all the conditions within all instructions. Each condition is repeated ten times.

| Instructions | Distance (% leg length) | Direction |
| --- | --- | --- |
| Standard | 45 | 0° |
| Standard | 45 | 45° |
| Standard | 80 | 0° |
| Standard | 80 | 45° |
| Fast | 45 | 0° |
| Fast | 45 | 45° |
| Fast | 80 | 0° |
| Fast | 80 | 45° |
| Accurate | 45 | 0° |
| Accurate | 45 | 45° |
| Accurate | 80 | 0° |
| Accurate | 80 | 45° |

This design enables the examination of how instructional emphasis, stimulus processing, movement planning, and execution interact during visually cued volitional stepping (Figure 3).

**Figure 3.**
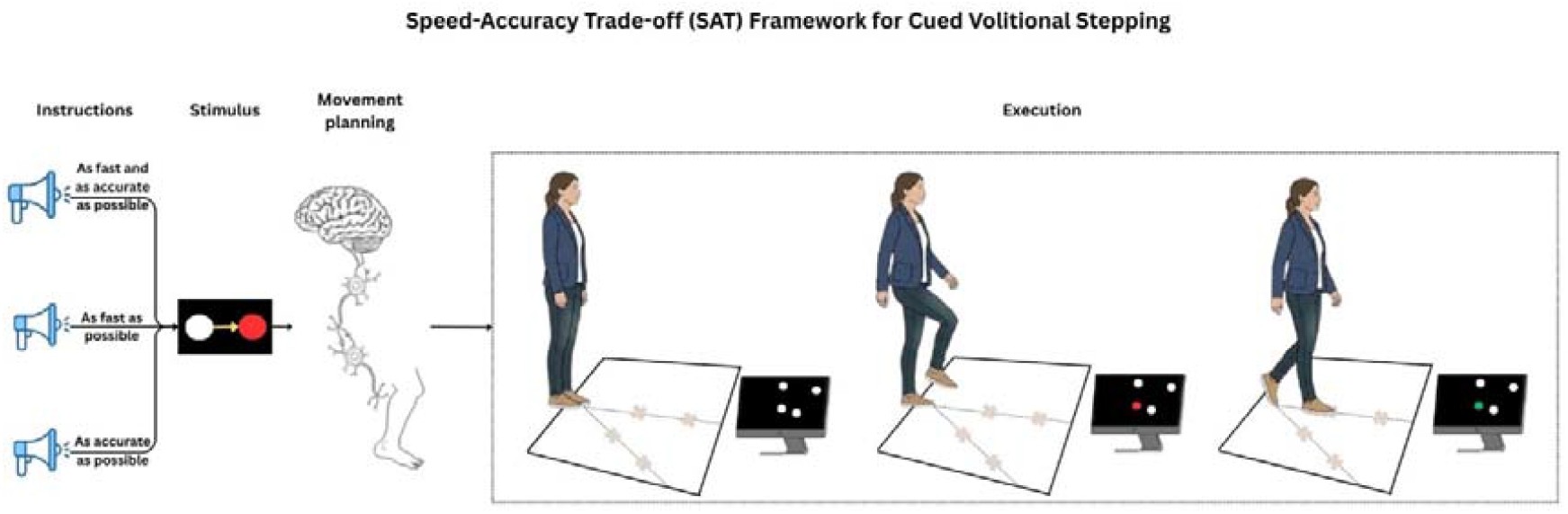
Stepping paradigm.

### Analysis Plan

#### Metrics

The metrics we will compute for each of the four targets and within each instruction block are (Figure 4):

**Figure 4.**
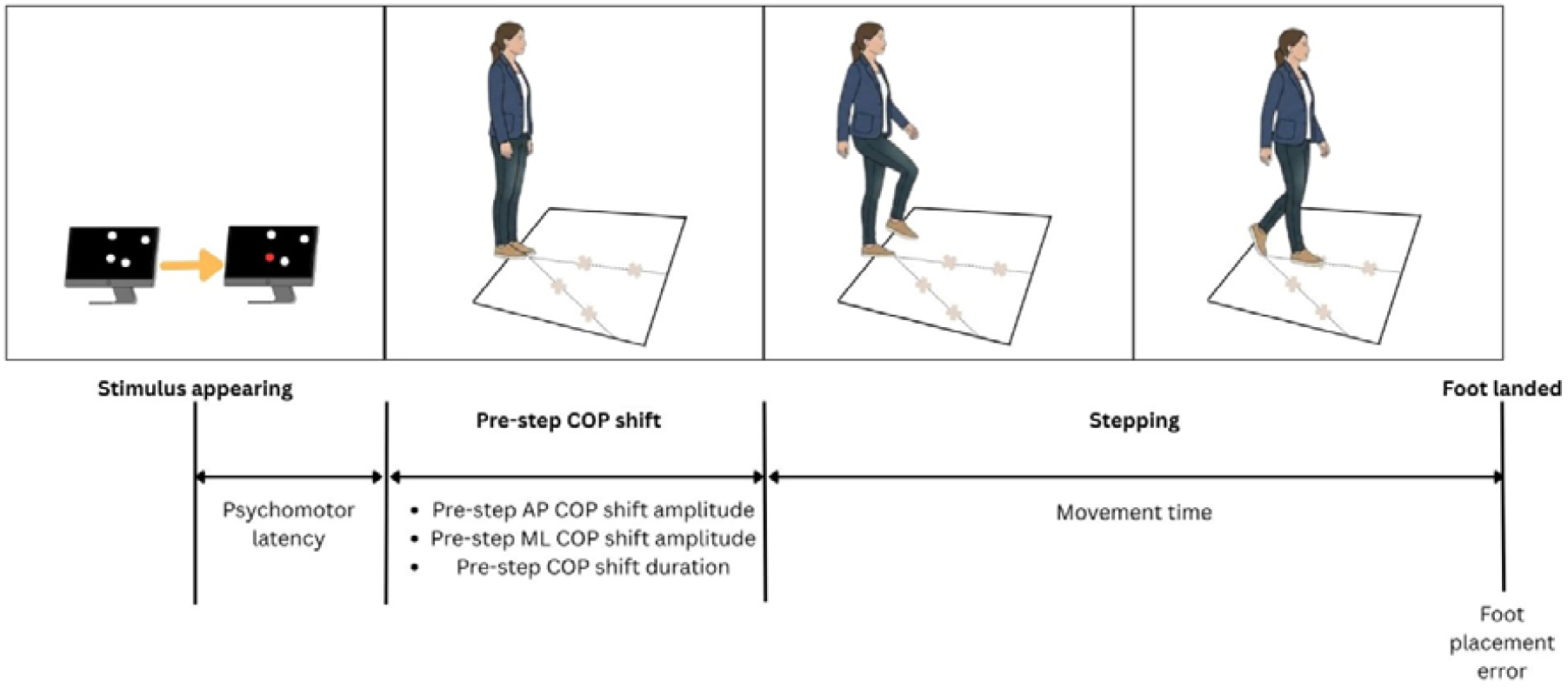
Schematic representation of the visually cued stepping paradigm and associated outcome measures.

- *Movement Time (MT)*: the duration between the onset of the leading foot’s movement and its landing on the target. Movement onset and end will be detected using a threshold-based algorithm on the peak velocity of the stepping HEEL and TOE, respectively, as commonly reported in previous studies (e.g., Danion et al., 1999; Duarte & Latash, 2007; Bertucco et al., 2010).
- *Foot placement error (FPE): the spatial deviation between the participant’s toe and the target location. The FPE for each trial will be calculated as the Euclidean distance between the toe at touchdown and the center of the target:*

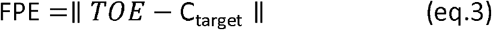

This approach is consistent with previous studies analyzing foot placement accuracy in standing and stepping tasks (e.g.,; Duarte & Latash, 2007; Bertucco et al., 2010).
- *Pre-step* mediolateral (ML) COP *shift amplitude*: maximum COP displacement along the ML axis prior to step initiation with respect to the initial position.
- Pre-step anteroposterior (AP) COP shift amplitude: maximum COP displacement along the AP axis prior to step initiation with respect to the initial position.
- *Pre-step COP shift duration*: the time interval between COP shift onset and step initiation, with COP shift onset detected using a threshold-based algorithm on the peak of COP velocity magnitude (e.g., Danion et al., 1999).
- *Psychomotor latency*: quantified as the time between stimulus presentation (i.e., dot coloring on the screen) and the pre-step COP shift onset.

### Statistical Analyses

#### Primary analysis

The primary analysis will address the speed-accuracy trade-off using two complementary, aggregate-level approaches for each condition: (i) a direct comparison between instructions (Fast vs. Accurate) to evaluate overall shifts in performance, and (ii) a modeling of the relationship between movement time and task difficulty, as well as foot placement error and task difficulty, using subject-level means

#### Speed vs accuracy

To assess the effect of prioritizing speed versus accuracy, comparisons will be restricted to the fast and accurate conditions.

For each participant, a mean value for each metric (MT, FPE, pre-step AP and ML COP shift amplitudes, pre-step COP shift duration, and psychomotor latency) will be calculated for every experimental condition. To determine the impact of instruction across different biomechanical demands, the Fast and Accurate instructions will be compared separately within each combination of direction (0°, 45°) and distance (45%, 80% leg length). Prior to these comparisons, the normality of the data distribution for each condition will be evaluated using the Shapiro-Wilk test. If assumptions of normality are met, paired-sample t-tests will be used; otherwise, the Wilcoxon signed-rank test will be applied. To account for multiple comparisons across the four conditions, a Bonferroni correction will be applied to the alpha level.

To further quantify the trade-off, individual difference scores (Δ) between the Fast and Accurate conditions (e.g., *ΔMT = MT*_*fast*_ − *MT*_*accurate*_) will be calculated for each metric. A series of correlation analyses will be performed on these delta scores to determine if the magnitude of change in movement speed (ΔMT) is significantly associated with the magnitude of change in stepping accuracy (ΔFPE) and postural preparation (ΔCOP parameters). Prior to these analyses, the distribution of the delta scores will be assessed for normality using the Shapiro-Wilk test. Pearson’s correlation coefficient (r) will be used if the data meet the assumptions of normality; otherwise, Spearman’s rank correlation (rho) will be employed. This approach will allow us to determine whether individual adjustments in movement speed directly correspond to systematic changes in accuracy and preparatory postural strategies.

#### Fitts law

To better account for variability in lower-limb tasks, an effective target width (W11) has been proposed, defined on the basis of endpoint variability:

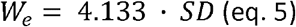

where SD is the standard deviation of the final landing positions across trials (Bertucco & Cesari, 2010), calculated as the Euclidean distance from the starting point to the toe at touchdown. This leads to an effective index of difficulty:

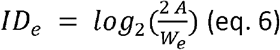

which captures functional difficulty by incorporating performance variability, making it particularly relevant for stepping tasks where motor variability and postural control are tightly coupled.

To account for task difficulty, the effective index of difficulty (*ID*_*e*_) will be computed as eq. 6. Separate linear regression models will be fitted for each movement direction (0° and 45°) to describe the relationships between movement time and task difficulty (eq. 7) and between foot placement error and task difficulty (eq. 8).

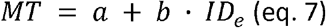

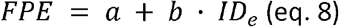

Where MT and FPE are the average across the ten trials for each condition.

The slope (*b*) will be interpreted as the rate of information processing, quantifying the marginal cost, in either time or spatial precision, added by each increase in task difficulty (*ID*_*e*_). In MT model (eq. 7), a steeper slope will indicate a lower processing efficiency, as the motor system requires more time to plan and execute a step for a given increase of ID. In the FPE model (eq. 8), the slope will represent the spatial error scaling factor, where a steeper slope will indicate that stepping accuracy is more sensitive to, and more easily degraded by, increasing task demands. Differences in slope between directions (0° vs. 45°) will be used to determine whether the additional biomechanical and postural requirements of diagonal stepping significantly tax the efficiency of the motor control system.

The fit of the regression models will be assessed using the Coefficient of Determination (*R*^2^). Residual analysis will be performed to verify model assumptions, including linearity, homoscedasticity, and absence of systematic bias, ensuring that the models adequately captures the relationships between task difficulty and movement time, and between task difficulty and foot placement error, respectively.

### Secondary analyses

The secondary analyses will examine the underlying granular structure of the data through three complementary, trial-level approaches: (i) Linear Mixed Modeling (LMM) to investigate the effects of instruction, direction, and distance while accounting for baseline individual differences across every repetitions; (ii) trial-wise Fitts’ law modeling to evaluate the robustness and scaling of the speed-accuracy trade-off at the single-trial level; and (iii) quantification of motor consistency through the analysis of intra-individual variability to assess neuromotor noise across conditions.

#### Effects of Instruction, Direction, and Position on Stepping Performance

To examine the effects of instruction, direction, and position while accounting for baseline individual differences, mean performance values for each participant (calculated across all trials within each condition) will be analyzed using Linear Mixed Models (LMM). Fixed effects will include Instruction (Fast, Accurate, Fast & Accurate), Direction (0°, 45°), and Position (45%, 80% leg length). To account for baseline reaction and postural capacities, Reaction Time and COP path length (obtained from baseline tests) will be included as continuous covariates in the models.

Separate models will be constructed for each dependent variable: MT, FPE, pre-step COP shift duration, and pre-step COP shift amplitudes (analyzed separately for ML and AP axes). Significant main effects and interactions will be explored using Bonferroni-corrected post-hoc comparisons. Normality of residuals will be assessed via visual inspection of Q-Q plots and the Shapiro-Wilk test.

#### Trial-Level Fitts’ Law Analysis

To further investigate the robustness of the speed-accuracy trade-off under intra-individual variability, the Fitts’ law relationship will also be examined at the single-trial level.

Trial-wise movement time and foot placement error will be modeled as a function of task difficulty (eq. 7, 8).

To account for repeated measures within participants, a mixed-effects modeling approach will be used, including participant as a random effect. Separate models will be fitted for each movement direction (0° and 45°).

Model fit will be evaluated using the Coefficient of Determination (*R*^2^) and residual analysis. Comparisons with the aggregated-data models (primary analysis) will be used to assess the influence of intra-individual variability on the stability of the MT–*ID*_*e*_ and FPE–*ID*_*e*_ relationships.

#### Intra-Individual Variability

Intra-individual variability in movement time and foot placement error will be quantified using the Coefficient of Variation (CV), calculated for each participant and condition. Differences in variability across conditions will be explored using repeated-measures analyses (ANOVA or Friedman test, depending on assumptions). Elevated variability will be interpreted as reflecting reduced motor consistency and increased neuromotor noise, particularly in conditions requiring greater postural control.

### Sample Size

To determine the required sample size, we reviewed the literature regarding instruction-based modulation of stepping performance under speed-accuracy trade-offs. While several studies have explored stepping under various conditions, only one study (Uemura et al., 2013) specifically manipulated instructions to investigate the speed-accuracy trade-off in a comparable paradigm.

In Uemura et al. (2013), instruction significantly affected step execution time, with faster performance under speed instruction (0.81 ± 0.08 s) compared to accuracy instruction (0.94 ± 0.08 s), corresponding to a mean within-participant difference of approximately 0.13 s.

A standardized effect size was derived directly from these values by dividing the mean difference by the pooled standard deviation (SDp ≈ 0.08s), following the conventional definition of Cohen’s d for mean differences (Cohen, 1988):

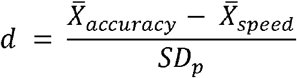

This yields an effect size of approximately d ≈ 1.6 for step execution time.

However, this estimate reflects a best-case scenario from a single outcome measure and may overestimate effects for a simple two-condition comparison. Therefore, a more conservative effect size of d = 0.8 was selected for power analysis of the primary comparison (fast vs accurate), consistent with typical effects reported in within-participant motor control paradigms.

A priori power analysis for a paired-sample t-test (α = 0.05, power = 0.95, two-tailed) indicated a required sample size of approximately 23 participants. To ensure balanced sex distribution, the final sample size was set to 24 participants.

## Code Availability Statement

The code used to process the raw marker trajectory and force plate data, as well as to perform the statistical analyses, will be made publicly available on GitHub.

## Data Availability Statement

The pseudonymized dataset will be published on Zenodo.

## Author contributions

Conceptualization: EG; Funding Acquisition: EG; Methodology: PSD, AIK, EG; Project Administration: PDF, AIK; Software: PDF; Resources: DKR, EG; Supervision: EG; Visualization: PDF, AIK; Writing-original draft: PDF, AIK; Writing-review & editing: PDF, AIK, WZ, DKR, OL, RG, EG.

## Competing interests

The authors declare no competing interests.

## Notes

### Competing Interest Statement

The authors have declared no competing interest.

### Author Declarations

The study protocol was approved by the ethics committees of ETH Zurich (Nr: 26 ETHICS-053).

## References

Altorfer, P., Adcock, M., de Bruin, E. D., Graf, F., & Giannouli, E. (2021). Feasibility of cognitive-motor exergames in geriatric inpatient rehabilitation: a pilot randomized controlled study. Frontiers in aging neuroscience, 13, 739948.

Bertucco, M., & Cesari, P. (2010). Does movement planning follow Fitts’ law? Scaling anticipatory postural adjustments with movement speed and accuracy. Neuroscience, 171(1), 205–213. 10.1016/j.neuroscience.2010.08.023

Bogacz, R., Wagenmakers, E.-J., Forstmann, B. U., & Nieuwenhuis, S. (2010). The neural basis of the speed–accuracy tradeoff. Trends in Neurosciences, 33(1), 10–16. 10.1016/j.tins.2009.09.002

Bradshaw, E. J., & Sparrow, W. A. (2000). The Speed-Accuracy Trade-Off in Human Gait Control When Running Towards Targets. Journal of Applied Biomechanics, 16(4), 331–341. 10.1123/jab.16.4.331

Büttiker, J., Marks, D., Hanke, M., Ludyga, S., Marsico, P., Eggimann, B., & Giannouli, E. (2024). Cognitive-motor exergame training on a labile surface in stroke inpatients: study protocol for a randomized controlled trial. Frontiers in Neurology, 15, 1402145.

Day, B. L., & Bancroft, M. J. (2018). Voluntary steps and gait initiation. Handbook of clinical neurology, 159, 107–118.

Duarte, M., & Latash, M. L. (2007). Effects of postural task requirements on the speed– accuracy trade-off. Experimental Brain Research, 180(3), 457–467. 10.1007/s00221-007-0871-x

Fitts, P. M. (1954). The information capacity of the human motor system in controlling the amplitude of movement.

Hayashi, H., Tomita, H., Ogawa, Y., Kawamata, N., Motomura, S., & Kanai, A. (2025). Anticipatory postural adjustments during multidirectional gait initiation with body turning among healthy young men. Gait & Posture, 110003.

Heitz, R. P. (2014). The speed-accuracy tradeoff: History, physiology, methodology, and behavior. Frontiers in Neuroscience, 8. 10.3389/fnins.2014.00150

Inaba, Y., Suzuki, T., Yoshioka, S., & Fukashiro, S. (2020). Directional control mechanisms in multidirectional step initiating tasks. Frontiers in human neuroscience, 14, 178.

Iwase, H., & Murata, A. (2001, September). Modelling of human’s three-dimensional movement-extending Fitts’ model to three-dimensional pointing task. In Proceedings 10th IEEE International Workshop on Robot and Human Interactive Communication. ROMAN 2001 (Cat. No. 01TH8591) (pp. 594–599). IEEE.

Jäggi, S., Wachter, A., Adcock, M., de Bruin, E. D., Möller, J. C., Marks, D., & Giannouli, E. (2023). Feasibility and effects of cognitive–motor exergames on fall risk factors in typical and atypical Parkinson’s inpatients: a randomized controlled pilot study. European journal of medical research, 28(1), 30.

Katsimpokis, D., Hawkins, G. E., & Van Maanen, L. (2020). Not all Speed-Accuracy Trade-Off Manipulations Have the Same Psychological Effect. Computational Brain & Behavior, 3(3), 252–268. 10.1007/s42113-020-00074-y

Lakens, D. (2013). Calculating and reporting effect sizes to facilitate cumulative science: a practical primer for t-tests and ANOVAs. Frontiers in psychology, 4, 62627.

Luchies, C. W., Schiffman, J., Richards, L. G., Thompson, M. R., Bazuin, D., & DeYoung, A. J. (2002). Effects of Age, Step Direction, and Reaction Condition on the Ability to Step Quickly. The Journals of Gerontology Series A: Biological Sciences and Medical Sciences, 57(4), M246–M249. 10.1093/gerona/57.4.M246

MacKenzie, I. S., & Buxton, W. (1992, June). Extending Fitts’ law to two-dimensional tasks. In Proceedings of the SIGCHI conference on Human factors in computing systems (pp. 219–226).

Massion, J. (1992). Movement, posture and equilibrium: Interaction and coordination. Progress in Neurobiology, 38(1), 35–56. 10.1016/0301-0082(92)90034-C

Muijres, W., Arnalsteen, S., Daenens, C., Afschrift, M., & De Groote, F. (2023). Accuracy-speed-stability trade-offs in a targeted stepping task are similar in young and older adults. Frontiers in Aging Neuroscience, 15, 1130707. 10.3389/fnagi.2023.1130707

Okubo, Y., Schoene, D., Caetano, M. J., Pliner, E. M., Osuka, Y., Toson, B., & Lord, S. R. (2021). Stepping impairment and falls in older adults: A systematic review and meta-analysis of volitional and reactive step tests. Ageing Research Reviews, 66, 101238. 10.1016/j.arr.2020.101238

Plamondon, R., & Alimi, A. M. (1997). Speed/accuracy trade-offs in target-directed movements. Behavioral and Brain Sciences, 20(2), 279–303. 10.1017/S0140525X97001441

Russo, Y., Marinkovic, D., Obradovic, B., & Vannozzi, G. (2021). Characterization of anticipatory postural adjustments in lateral stepping: impact of footwear and lower limb preference. Sensors, 21(24), 8244.

Schmidt, R. A., Lee, T. D., Winstein, C. J., Wulf, G., & Zelaznik, H. N. (2019). Motor control and learning: A behavioral emphasis (Sixth edition). Human Kinetics.

Seinsche, J., de Bruin, E. D., Saibene, E., Rizzo, F., Carpinella, I., Ferrarin, M., & Giannouli, E. (2023). Feasibility and effectiveness of a personalized home-based motor-cognitive training program in community-dwelling older adults: protocol for a pragmatic pilot randomized controlled trial. JMIR Research Protocols, 12(1), e49377.

Uemura, K., Oya, T., & Uchiyama, Y. (2013). Effects of speed and accuracy strategy on choice step execution in response to the flanker interference task. Human Movement Science, 32(6), 1393–1403. 10.1016/j.humov.2013.07.007

van Melick, N., Meddeler, B. M., Hoogeboom, T. J., Nijhuis-van der Sanden, M. W., & van Cingel, R. E. (2017). How to determine leg dominance: The agreement between self-reported and observed performance in healthy adults. PloS one, 12(12), e0189876.

